# Limitations of cross-border containment strategies for Bundibugyo ebolavirus

**DOI:** 10.64898/2026.06.04.26354820

**Authors:** Casey E. Middleton, Daniel B. Larremore

**Affiliations:** Department of Computer Science, University of Colorado Boulder, Boulder, CO 80309 USA; BioFrontiers Institute, University of Colorado Boulder, Boulder, CO 80303 USA; Santa Fe Institute, Santa Fe, NM 87501 USA

## Abstract

An ongoing outbreak of Bundibugyo virus disease (BVD) in the Democratic Republic of the Congo was deemed a public health emergency of international concern in May 2026. To prevent cross-border importation, many countries have already proposed containment strategies, and others are likely to follow suit. How well (or poorly) are screening and quarantine containment measures are likely to work for BVD? We leverage established epidemiological theory and develop a mathematical model of traveler screening and post-arrival quarantine to answer this question. We find that traveler screening via symptom screening or molecular testing will miss the majority of infected travelers, and should be complemented by post-arrival quarantine and monitoring of sufficient duration to detect those with long incubation periods. Our findings underscore the limitations of border screening and the importance of complementary measures like post-arrival quarantine to prevent local importation of BVD.

---

An ongoing outbreak of Bundibugyo virus disease (BVD), an Ebola disease caused by the *Bundibugyo ebolavirus*, was deemed a public health emergency of international concern in May 2026 [1]. To mitigate cross-border importation risk, numerous countries, including the United States, Canada, India, Thailand, and Kenya have already proposed travel bans, screening measures, and post-arrival quarantine [2, 3], and others are likely to follow suit. However, cross-border containment strategies are subject to a number of limitations, which we outline here.

Traveler screening has previously been implemented to prevent cross-border transmission of Ebola virus [4], but there are three fundamental challenges that limit the effectiveness of screening travelers. First, the effectiveness of symptom-based screening is fundamentally limited by the incubation period. Even under optimistic sensitivity scenarios, symptom-based screening misses the majority of cases because individuals travel during their presymptomatic incubation period [5]. Second, the effectiveness of screening with molecular tests is fundamentally limited by the gap between infection and detectability, the days or weeks after exposure during which target molecules are below the limits of detection of even the most sensitive diagnostics [6]. Finally, the challenges faced by symptom screening and molecular tests are compounded during a growing epidemic. In exactly the same way that a growing human population skews demographically younger, a growing epidemic’s infections skew toward more recent infections [7]—precisely those that expose the two weaknesses of symptom and diagnostic screening strategies above. Moreover, the infections missed by these traveler screening programs are those whose potential to infect others lies ahead of them, making them more likely to seed an epidemic after arrival at their destination [6].

In light of these challenges, travel bans have been enacted to limit the number of potentially infected travelers [3]. While travel bans were shown to reduce importation by a few days to weeks during the 2014 Ebola epidemic [8], they may have also prevented the delivery of emergency supplies and responders [4]. Alternatively, post-arrival interventions, such as symptom monitoring, diagnostic testing, and quarantine, could detect those who are missed by traveler screening. However, the impact of post-arrival monitoring depends on the duration of monitoring being sufficient to detect all infections and the ability of monitoring to trigger effective interventions when needed — and programs with a sufficient quarantine duration to catch even the longest incubation periods are extremely costly.

## Expected impact of cross-border containment strategies for Bundibugyo virus

While epidemiological theory can support understanding of the general limitations of cross-border containment strategies, past BVD outbreaks can inform quantitative predictions specific to the current epidemic. The average incubation period for BVD is estimated to be 6.3d (95% confidence interval, 5.2d–7.3d; [9]), meaning that pre-symptomatic cases may travel unknowingly for nearly a week after exposure. Under these conditions, how effective is screening predicted to be?

We developed a simple model to predict the proportion of infected travelers missed by symptom screening at arrival and post-arrival quarantine with symptom monitoring (Supplemental Materials). In this model, we omitted screening with molecular tests due to the limited availability of BVD-specific tests at the time of writing, but previous estimates show that even using high sensitivity Ebola virus tests for traveler screening miss 89.5% of infected travelers [6].

Assuming that infected travelers become too sick to travel 1-3 days after symptom onset [10], our model estimates that arrival symptom screening will miss 67.6%–86.3% of infected travelers in a steady state outbreak within the source country (Supp. Table 1). Symptom screening will miss a smaller proportion of infections if severity never prohibits travel and misses more during a growing epidemic (Supp. Figure 1, Supp. Table 1).

If all arriving travelers are subject to a universal post-arrival quarantine and monitoring, we estimate that a 7d quarantine will miss 26.3% of infections. If both interventions are applied, the combination of arrival screening with post-arrival universal quarantine is predicted to miss 17.8%–22.7% of infected travelers. In other words, we predict a slight improvement from traveler screening alone, but the quarantine does most of the work. Increasing quarantine duration decreases the proportion of infections missed, but even a 21-day quarantine is predicted to be imperfect if the incubation period distribution is heavy-tailed (Supp. Figure 1), though empirical data to better characterize the shape of this distribution would greatly improve predictions. Thus, quarantine decisions must weigh the increasing cost of longer quarantine periods against the diminishing benefit of reduced risk of evasion.

In light of our evolving understanding of BVD, our analysis shows that post-arrival quarantine measures will play a more important role in preventing importation than symptom screening. Increased availability of high sensitivity molecular tests may reduce the number of travelers missed by traveler screening and the duration of post-arrival quarantine required to confidently rule out infection, but molecular tests will not eliminate the need for post-arrival quarantine. Improved understanding of the incubation period distribution and time-varying sensitivity of molecular assays would help us better inform interventions critical to containment.

Thus, when considering the efforts-to-risk tradeoffs of various cross-border containment strategies, policy-makers should consider that increasing the duration of post-arrival quarantine and monitoring may be a more effective intervention than traveler screening.

## Data Availability

All results from this study are reproducible from the mathematical model and parameters provided in the Supplemental Materials. Code is available from the authors upon request.

## Supplemental Materials

### Model

Consider a single infected individual. Let *X* be a random variable describing the individual’s incubation period, with probability density function (PDF) *f* (*x*). Let *t* denote the infection age at the time of travel, representing the number of days since infection at the moment an individual attempts to travel. Let *q*(*t*) and *Q*(*t*) define the PDF and CDF, respectively, for the infection age at which an individual from the general population attempts travel.

A symptom-based screening program at the border catches travelers who are visibly symptomatic. However, Ebola symptoms typically progress from mild, non-specific illness to more severe illness within the first few days of disease [10], and the progression to severe illness may itself inhibit travel. Thus, an individual with incubation period *X* who attempts travel at infection age *t* falls into one of three categories:

- *t < X*: The traveler is still pre-symptomatic and slips through screening undetected.
- *X ≤ t < X* + *α*: The traveler is symptomatic but not yet too ill to travel, and is caught by screening.
- *t ≥ X* + *α*: The traveler is too sick to travel and never attempts the trip.

The parameter *α* denotes the window of time between symptom onset and severe physical incapacitation. We consider scenarios where symptom severity prevents travel 1–3 days after illness onset [10], or a scenario where illness never prevents travel (*α* = *∞*).

Because individuals do not attempt travel if *t ≥ X* + *α*, the distribution of incubation periods among actual travelers differs from that of the general population. Let *X*_trav_ be the random variable for the incubation period of an individual who successfully attempts travel (i.e., conditioned on *t < X* + *α*). By Bayes’ theorem, the PDF of *X*_trav_ is given by:

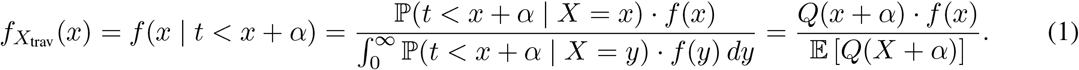

#### Arrival Symptom Screening

Let *ρ* denote the failure rate of the screening program, defined as the probability that an infected traveler who attempts a trip slips through undetected. For an individual traveler with a realized incubation period *x*, they evade detection if they travel while pre-symptomatic (*t < x*), given that they are able to travel at all (*t < x* + *α*). This conditional probability is:

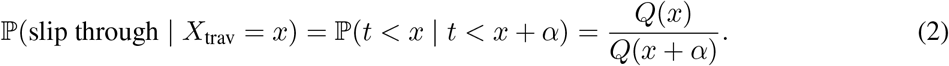

To find the overall failure rate *ρ*, we take the expectation of this probability over the incubation period distribution of actual travelers, *X*_trav_:

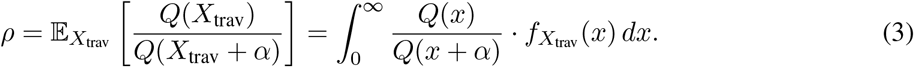

Substituting the definition of 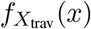 into this integral yields:

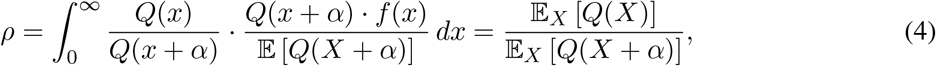

where the expectations in the final fraction are now taken over the general population distribution, *X*.

##### Uniform infection ages

In a steady-state outbreak, the infection age at travel is uniformly distributed across the infected population (*Q*(*t*)*∝ t*). Under this assumption, the failure rate simplifies directly to a ratio of the expected timelines:

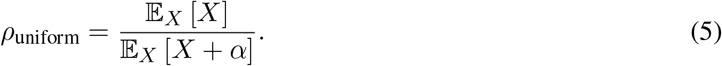

##### Non-uniform infection ages

When the epidemic is growing exponentially at rate *r*, “younger” infections outnumber “older” ones due to expanding incidence [7]. A traveler attempting travel today at infection age *t* was infected *t* time units ago, when population incidence was proportional to *e*^*−rt*^. This shifts the infection-age distributions to:

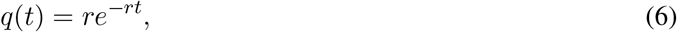

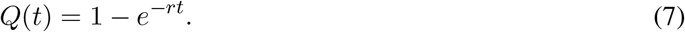

Substituting *Q*(*t*) into the general failure rate derivation yields:

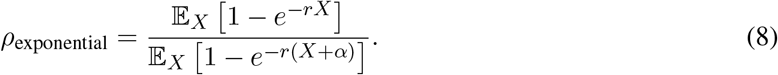

### Quarantine at Arrival

Consider an infected person with incubation period *X* and a potential travel infection age *t*. Suppose the destination country imposes a mandatory quarantine of duration *b* upon arrival.

To slip through the quarantine program undetected, the traveler must arrive pre-symptomatic (*t < X*) and remain asymptomatic for the entire duration of the quarantine window, meaning their infection age at quarantine exit is less than their incubation period (*t* + *b < X*).

For a traveler with a realized incubation period *x*, the probability of evading detection given that they successfully attempted travel (*t < x* + *α*) is:

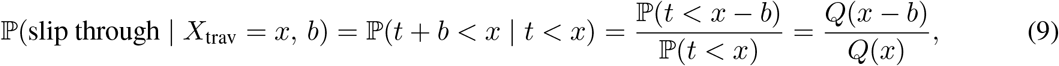

where *Q*(*x − b*) = 0 for all *x ≤ b*, reflecting the reality that individuals with incubation periods shorter than the quarantine duration will always develop symptoms before release.

To find the overall failure rate of the quarantine program, we take the expectation of this conditional probability over the incubation period distribution of actual travelers, *X*_trav_:

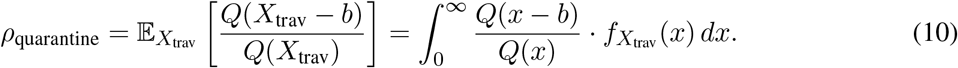

Substituting the traveler PDF, 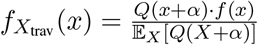, into the integral yields:

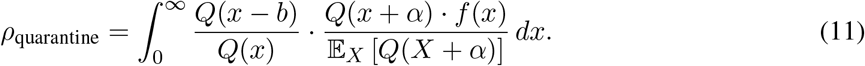

Notice that if we evaluate this under the baseline assumption that arrival symptom screening occurs right before quarantine entry, the probability of an infected individual successfully boarding and slipping through the initial arrival screen is exactly governed by the 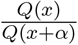 scaling we derived previously. Jointly evaluating entry screening and quarantine tracking allows the expectations to simplify cleanly over the general population distribution, *X*:

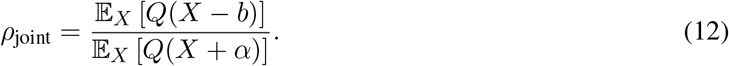

#### Incubation period distribution and parameter estimation for Bundibugyo virus

Following convention, we model incubation times using a lognormal distribution. Empirical data supporting or refuting this particular shape for the incubation period distribution would naturally improve this paper’s estimates.

To parameterize the lognormal distribution, we estimated mean and standard deviation parameters from published estimates as follows: MacNeil et al report a mean incubation period of *x* = 6.3 days with a 95% confidence interval of the mean of [5.2, 7.3] days for BVD, estimated from *n* = 56 observations. Assuming that MacNeil et al report their confidence interval of the mean using Normal statistics, we estimate the empirical standard deviation by asking what *σ*^2^ would have produced this CI with that sample size: 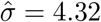 days.

**Table 1:**
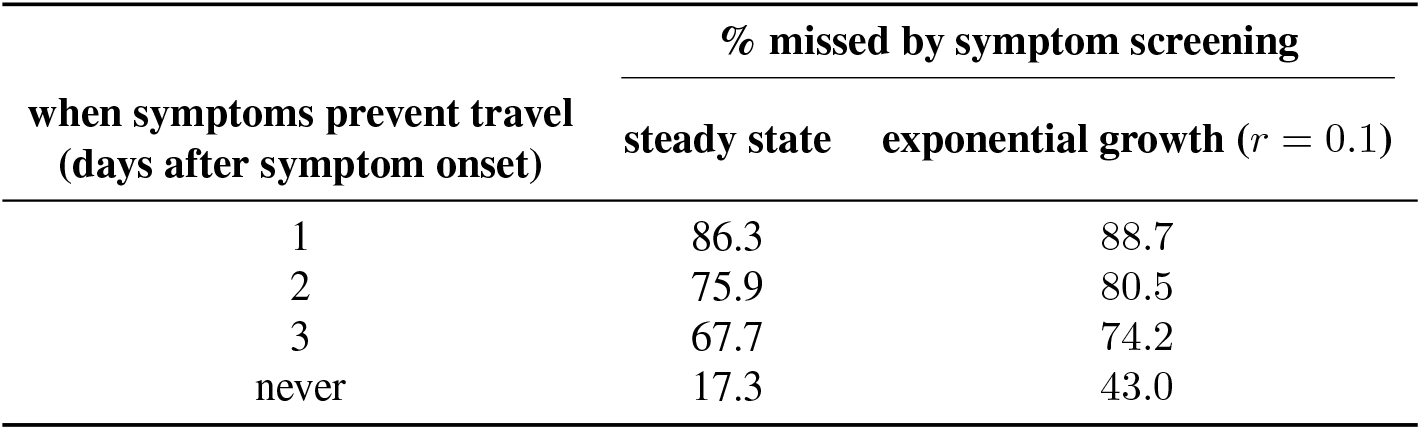
Percentage of infected individuals missed by arrival symptom screening under different epidemiological dynamics. Percentages are shown under steady state (i.e., endemic) and exponential growth (*r* = 0.1) scenarios at the source location, assuming that symptom severity prevents travel 1-3 days after onset or never.

**Figure 1:**
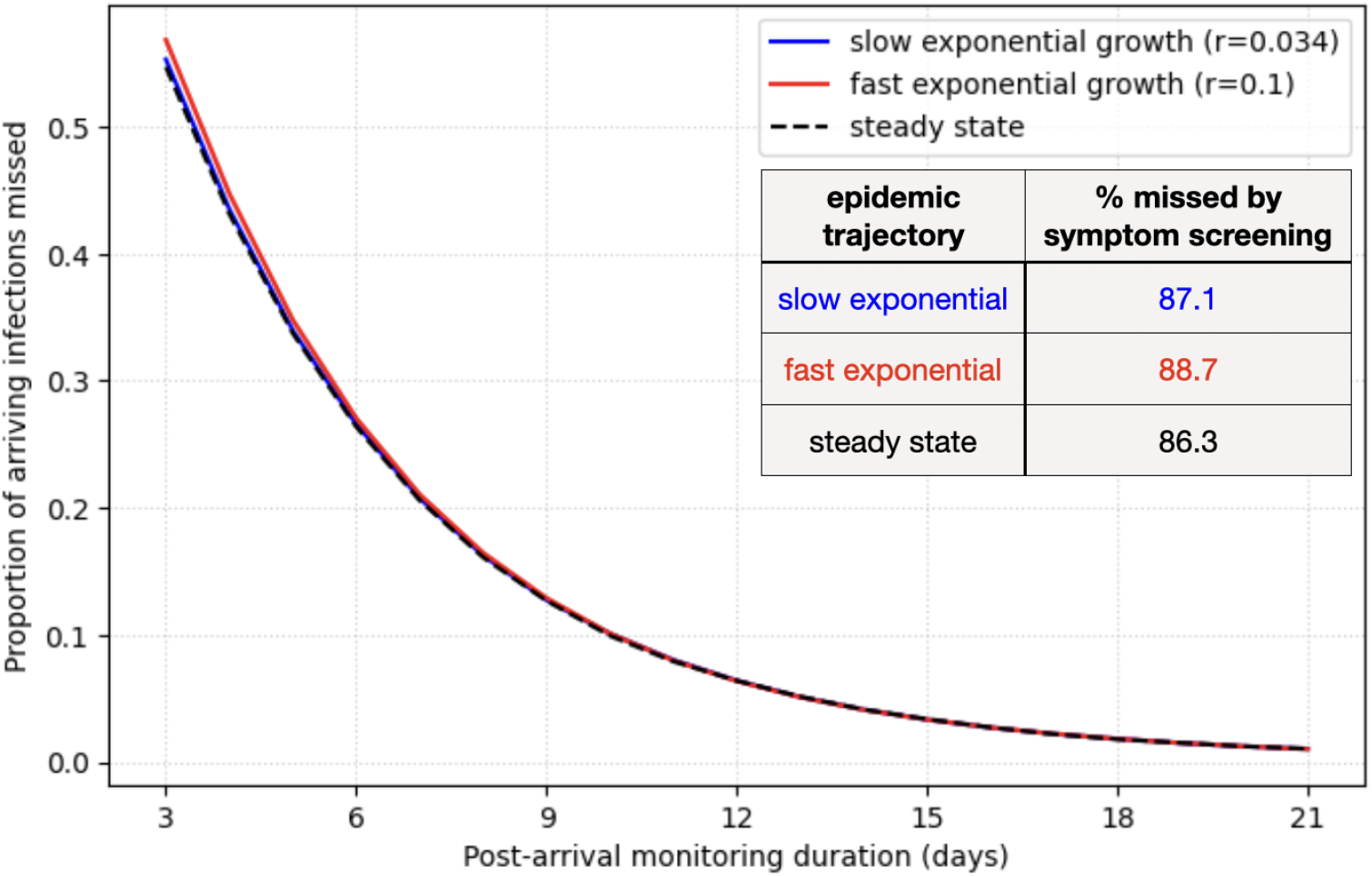
Effectiveness of pre-departure symptom screening and post-arrival quarantine is dominated by post-arrival quarantine. The proportion of cases missed by symptom screening (table) and post-arrival quarantine (plot) are shown for two exponential growth scenarios and one steady state outbreak scenario. All scenarios assume an incubation period drawn from a lognormal distribution with a mean of 6.3 days and a standard deviation of 4.32 days.

